# Long COVID in people with multiple sclerosis and related disorders: a multicenter cross-sectional study

**DOI:** 10.1101/2024.12.20.24319365

**Authors:** Chen Hu, Jiyeon Son, Lindsay McAlpine, Elizabeth LS Walker, Megan Dahl, Emily Song, Sugeidy Ferreira Brito, Katelyn Kavak, Kaho Onomichi, Amit Bar-Or, Christopher Perrone, Claire S. Riley, Bianca Weinstock Guttman, Philip L. De Jager, Erin E Longbrake, Zongqi Xia

## Abstract

**Background:** Managing long COVID in people with multiple sclerosis and related disorders (pwMSRD) is complex due to overlapping symptoms. To address evidence gaps, we evaluated long COVID susceptibility in pwMSRD versus controls and its associations with multi-domain function and disability.

**Methods:** In this multicenter cross-sectional study, participants completed a survey covering 71 post-infection symptoms, distinguishing new-onset from worsening symptoms. We defined long COVID using the 2024 NASEM criteria. Logistic regression assessed long COVID odds. Linear and Poisson regression evaluated associations with function and disability.

**Results:** 969 pwMSRD (82.5% female, mean age 51.8 years, 63.5% infected) and 1,003 controls (79.4% female, mean age 45.2 years, 61.2% infected) were included. PwMSRD had higher odds of long COVID (aOR=1.6 [1.2-2.1]), with a stronger association when restricting to worsening symptoms (aOR=2.3 [1.7-3.1]). Having long COVID was associated with worse physical function, cognition, and depression in both groups. PwMSRD with long COVID experienced greater physical function declines and more depression severity exacerbation than controls, and had faster disability progression compared to those without long COVID.

**Conclusion:** PwMSRD show increased susceptibility to long COVID, primarily driven by worsening symptoms. Long COVID contributes to more functional decline and disability worsening. Recognizing and managing long COVID is essential in pwMSRD.

## Introduction

The COVID-19 pandemic has disproportionately affected individuals with multiple sclerosis and related disorders (pwMSRD).^1^ However, the pathophysiology and clinical manifestation of long COVID, also known as post-acute sequelae of COVID-19 (PASC), is poorly understood in this population.^2^ Long COVID has been widely recognized as a major public health concern in the general population, while few studies have examined its epidemiology or its impact on disability among pwMSRD. As a result, it remains unclear whether pwMSRD are more susceptible to long COVID and whether long COVID exacerbates the adverse effects of neuroinflammatory diseases on health outcomes.^3–8^

A variety of terms and definitions have been proposed for long COVID, each with distinct strengths and limitations. In 2023, the National Institutes of Health’s Researching COVID to Enhance Recovery (NIH RECOVER) initiative introduced a stringent definition, using a weighted scoring system based on 12 symptoms to identify PASC positivity.^6^ Although this approach enhances specificity, it is limited by selection bias, low sensitivity, and a lack of external validation among other populations. In 2024, the National Academies of Sciences, Engineering, and Medicine (NASEM) released a consensus definition of long COVID as “an infection-associated chronic condition occurring after SARS-CoV-2 infection, persisting for at least three months in a continuous, relapsing-remitting, or progressive manner, and affecting one or more organ systems.”.^9^ While intentionally inclusive, this definition lacks specificity due to the absence of symptoms or organ system specification.

In pwMSRD, long COVID is particularly challenging to characterize due to overlapping symptoms such as cognitive impairment, fatigue, and psychiatric manifestations.^10^ This overlap complicates the distinction between post-viral sequelae and underlying neuroinflammatory disease. Recent multimodal proteomic studies using machine learning approaches have suggested that active long COVID involves dysregulated complement activation, elevated antibody responses to herpesviruses, and thrombo-inflammatory markers, some of which are implicated in MS pathogenesis.^11^ Estimates of post-infectious symptom prevalence in pwMSRD range from 12.4% to 36.9%, but prior studies have not differentiated new symptoms from pre-existing ones.^10,12–14^ This distinction is crucial for diagnosing and managing long COVID in pwMSRD, and guiding treatment priorities. Finally, while long COVID has been shown to substantially impair patient-reported outcomes (PROs) in the general population, its specific burden in pwMSRD remains unquantified.^2,15^

The study objectives were to evaluate long COVID and PROs in a large, multi-center cohort of pwMSRD and controls. We distinguished new symptoms from worsening pre-existing symptoms to compare how post-infectious symptoms manifest in pwMSRD versus controls. We hypothesized that pwMSRD had increased odds of long COVID compared to controls and that both MSRD and long COVID independently contributed to worse PROs, with their combined presence leading to even greater impairment.

## Methods

### Study design and participants

This cross-sectional study utilized data from the Multiple Sclerosis Resilience to COVID-19 (MSReCOV) Collaborative, a cohort consisting of individuals with MSRD and controls, recruited from five U.S.-based clinical neuroimmunology centers (**Figure 1**).^16,17^ Participants enrolled in MSReCOV between April 2020 and July 2021. The MSRD group included individuals diagnosed with multiple sclerosis (MS), neuromyelitis optica spectrum disorder (NMOSD), myelin oligodendrocyte glycoprotein antibody-associated disease (MOGAD), and other rare neuroimmunological disorders (NID). The control group had no diagnosis of neuroinflammatory disorders, comprising relatives of pwMSRD, controls from local registries, and individuals recruited through local advertising at each participating center. Inclusion criteria for both groups were: (1) age ≥18 years; (2) able to provide informed consent; and (3) proficiency in English.^18,19^ Between August and December 2022, we invited all MSReCOV participants to complete a one-time survey via a centralized, secure, web-based Research Electronic Data Capture (REDCap) platform. Each participant completed the informed consent before starting the survey. The University of Pittsburgh Institutional Review Board approved the survey study (STUDY22080007).

**Figure 1.**
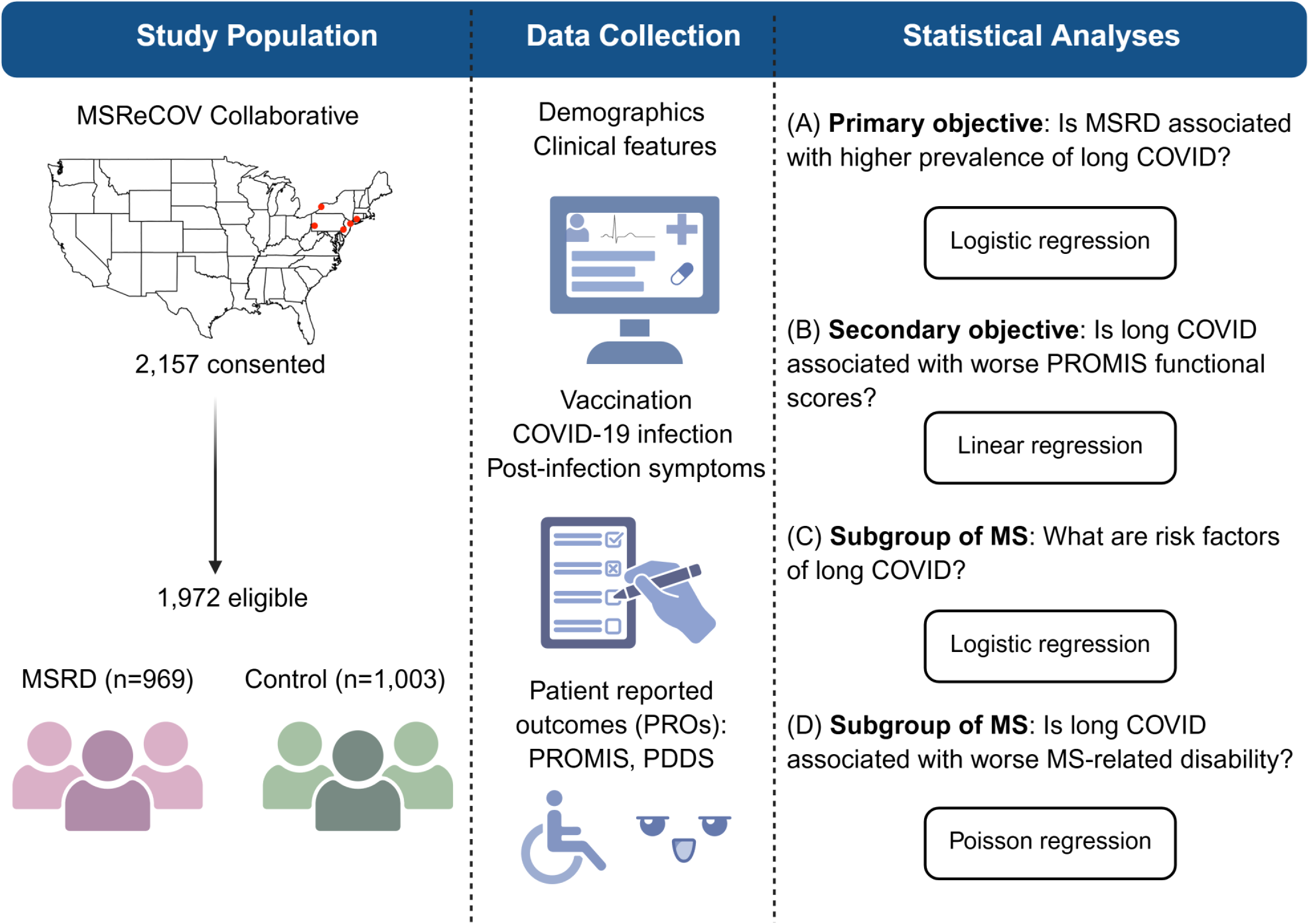
Study design. MSReCOV, Multiple Sclerosis Resilience to COVID-19; MSRD, Multiple sclerosis and related disorders; PROMIS, Patient-Reported Outcomes Measurement Information System; PDDS, Patient Determined Disease Steps.

### Measurements

#### Demographic and clinical profile

The survey collected information on age, sex, race, ethnicity, comorbidities adapted from the Charlson Comorbidity Index (CCI), height, weight, neuroimmunological diagnosis, and pre-pandemic employment status. Body mass index (BMI) was calculated using height and weight. Comorbidity burden was categorized as none (CCI=0), mild (1≤CCI≤ 2), moderate (3≤CCI≤ 5), or severe (CCI≥6). Employment status was classified as employed (including full-time, part-time, self-employed, students, and military personnel) or unemployed (including those out of work, retired, or unable to work). For pwMS, additional data were collected on MS subtype, classified as relapsing-remitting MS (RRMS) or progressive MS (including primary and secondary progressive type), and disease-modifying therapy (DMT) at survey, categorized into B-cell depletion therapies, sphingosine-1-phosphate (S1P) receptor modulators, other DMTs, or none/not answered, based on their prior associations with COVID-19 severity.^20^

#### COVID-19 infection and vaccine

Participants reported details regarding their COVID-19 infection history, including method of detection, dates of initial and subsequent infections, and severity of the initial infection. Acute COVID-19 infection was defined as having a confirmed or suspected infection based on a polymerase chain reaction (PCR) test, a rapid antigen test (either in a laboratory, at home, or in an uncertain setting), or self-reported symptoms consistent with the Centers for Disease Control and Prevention (CDC) list of COVID-19 symptoms. COVID-19 severity was categorized as asymptomatic, mild (symptomatic but able to function), moderate (symptomatic and very ill without hospitalization), severe (requiring hospital admission), or critical (requiring intensive care unit management). To account for variant-specific differences, the time of initial infection was classified as pre-Omicron (before January 1, 2022) or Omicron era (on or after January 1, 2022). Vaccination status at survey was categorized as fully vaccinated (receiving ≥3 doses) or not fully vaccinated (receiving <3 doses).

#### Long COVID symptoms

The primary study objective was to compare the odds of long COVID between pwMSRD and controls. Since no gold-standard symptom identification for long COVID exists (during study design), an expert panel of neurologists (JS, LM, PLD, EL, ZX) specializing in neuroimmunology developed a comprehensive symptom list, classifying 71 symptoms across 12 organ systems. To enhance validity, the questionnaire was adapted from the latest available knowledge of post-infection syndrome by organ systems at the time of survey deployment.^8^ Participants were asked whether they had experienced any of these symptoms occurring one month or later after their first acute COVID-19 episode, whether these symptoms were new or worsened compared to their pre-COVID baseline, and duration (months) of symptom persisted.

To define long COVID, we adopted the 2024 NASEM Long COVID definition for primary analyses.^9^ Participants who reported any symptoms that persisted for at least 3 months were classified as the long COVID group. To distinguish between long COVID driven by new symptoms and resulting from the worsening of pre-existing symptoms, we further restricted the long COVID status to new symptoms (i.e., absent during pre-COVID baseline) and worsening symptoms (i.e., worsened from the pre-COVID baseline). Based on these definitions, the three primary outcomes were overall long COVID (Yes vs No), long COVID based on new symptoms (Yes vs No), and long COVID based on worsening symptoms (Yes vs No).

#### Patient-reported outcomes

To assess functional outcomes, we utilized the Patient-Reported Outcomes Measurement Information System (PROMIS), including assessments of physical function (version 1.2), cognitive function (version 2.0), and depression (version 1.0). PROMIS instruments were standardized measures of patient-reported outcomes validated across various diseases. PROMIS T-scores were standardized to the U.S. general population, with a mean of 50 and a standard deviation of 10, with higher scores indicating greater levels of the measured attribute (e.g., higher depression scores indicated greater depressive severity). For pwMS, we additionally evaluated neurological disability both pre-COVID and at the time of survey using the Patient Determined Disease Steps (PDDS) scale, an ordinal scale ranging from 0 (no impairment) to 8 (bed-bound status).

Statistical analysis

The dataset included MSReCOV participants who completed the survey and had complete demographic, vaccination, and COVID-19 data. Participant characteristics were summarized as mean (SD), median (IQR), or frequency (proportion). Differences between pwMSRD and controls were assessed using the Wilcoxon rank-sum test for continuous variables and Fisher’s exact test for categorical variables.

Among participants with a history of acute COVID-19, we first compared the prevalence of each post-infection symptom between pwMSRD and controls using multivariable logistic regression, adjusting for age, sex, race/ethnicity, BMI, comorbidity burden (CCI), pre-pandemic employment, related factors of initial infection (i.e., detection method, severity, wave, and time to survey completion), reinfection, and vaccination status. We applied separate models for new-onset and worsening symptoms. A Bonferroni correction for 71 symptoms set the significance threshold at p<.0007. We then integrated individual symptoms into a composite outcome to define long COVID status using the NASEM definition and assessed three outcomes: overall long COVID, long COVID based on new symptom, and long COVID based on worsening symptom. We used logistic regression models to compare the odds of long COVID between pwMSRD and controls, adjusting for the same covariates.

To assess associations between long COVID status and PROs, we used multivariable linear regression models, further adjusting for MSRD status and its interaction with long COVID status to assess potential synergistic effects on health outcomes. Separate models examined physical function, cognition, and depression, using overall long COVID, long COVID based on new symptoms, and long COVID based on worsening symptoms as exposures. Within each model, we reported adjusted beta estimates and 95% CIs for long COVID status, MSRD status and their interaction.

In the subgroup of pwMS, we used multivariable logistic regression to identify factors associated with long COVID, including demographic and clinical profile (age, sex, race/ethnicity, BMI, comorbidities, employment), acute COVID-19 factors (detection method, wave, severity, reinfection, vaccination, time from infection to survey), and MS-specific characteristics (pre-COVID disability, MS subtype, DMT use). Significant factors were selected based on likelihood ratio tests (LRT) comparing full and reduced models. Finally, we employed Poisson regression to assess the relationship between long COVID and patient-reported disability based on PDDS, adjusting for the same set of demographic and clinical profile, acute COVID-19 features, and MS-specific characteristics. We reported adjusted rate ratios (aRRs) and 95% CIs of long COVID-associated disability worsening.

### Sensitivity analysis

The NASEM long COVID definition is limited in specificity. To test whether findings were robust to long COVID definitions, we alternatively applied the RECOVER scoring system, which classified the condition by assigning weighted points to each of symptom.^6^ Symptoms and their respective scores included loss of smell or taste (8 points), post-exertional malaise (7 points), chronic cough (4 points), brain fog (3 points), thirst (3 points), palpitations (2 points), chest pain (2 points), fatigue (1 point), sexual dysfunction (1 point), dizziness (1 point), gastrointestinal symptoms (1 point), abnormal movements (1 point), and hair loss (1 point). Participants with a total score of 12 or higher were classified as long COVID according to RECOVER.^6,10^ A list of the original symptoms surveyed and their classification within the RECOVER scoring system were provided in **eTable 1**. We separately computed scores for new symptoms and worsening symptoms, applying the same scoring thresholds. We reported the prevalence of RECOVER-defined long COVID, and its association with MSRD groups and PROs, using the same models employed in analyses based on NASEM-defined long COVID.

## Results

### Participants

Among the 3,527 MSReCOV participants invited to participate in the one-time survey, 2,157 consented. 1,972 participants (969 pwMSRD and 1,003 controls) completed the survey ≥3 months after the initial acute infection (**Figure 2)**. The consent rate was 61.2% (2,157/3,527), and the response rate among consented participants was 92% (1,972/2,157).

**Figure 2.**
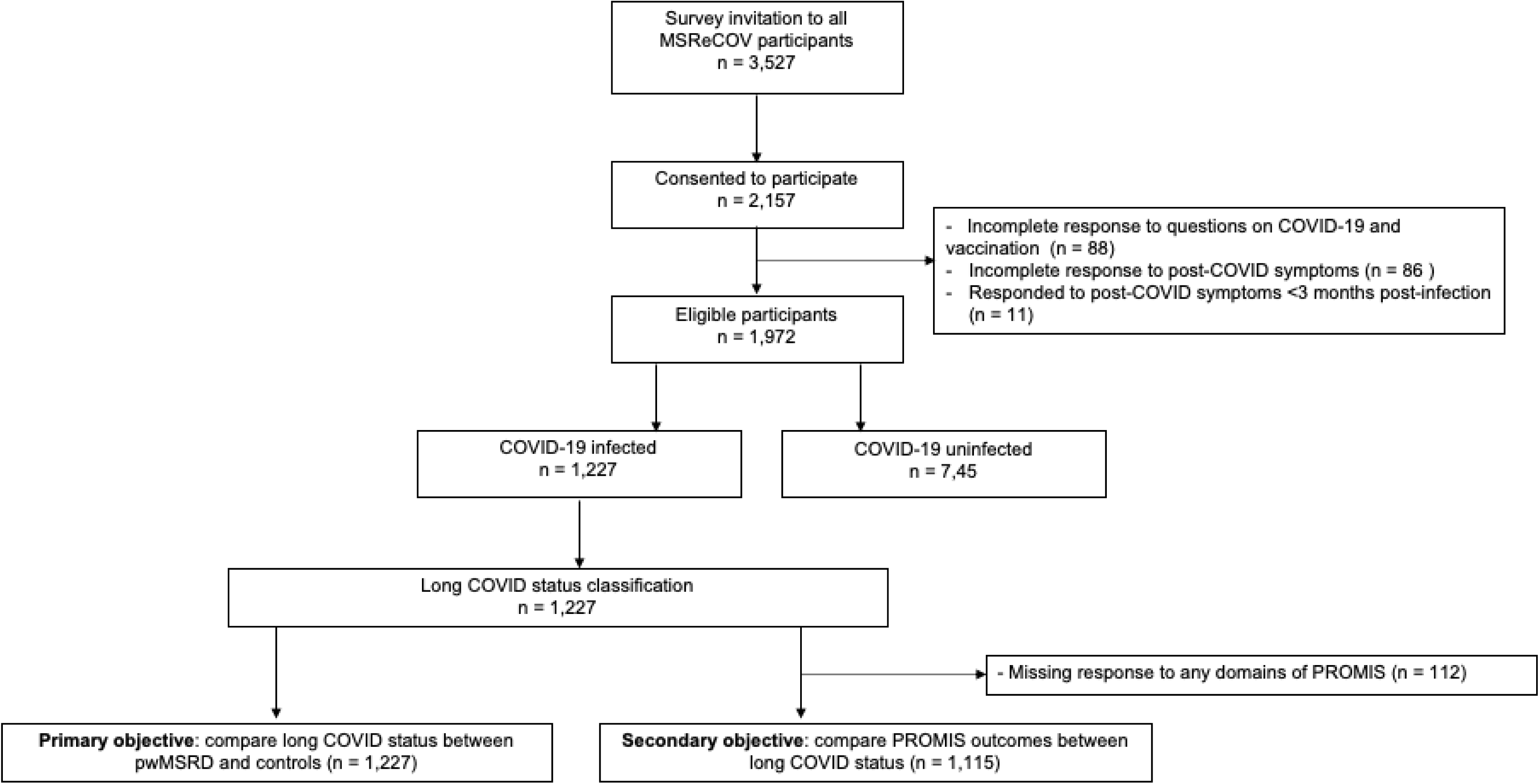
Participant disposition. MSReCOV, Multiple Sclerosis Resilience to COVID-19; MSRD, Multiple sclerosis and related disorders; PROMIS, Patient-Reported Outcomes Measurement Information System; pwMSRD, People with multiple sclerosis and related disorders.

Compared to survey responders, non-responders were younger on average, more likely to be pwMSRD, and less likely to be female or non-Hispanic White (**eTable 2**). When comparing pwMSRD with controls among participants who completed the survey (**Table 1**), pwMSRD were older (mean age 51.8 years [SD 12.1] vs 45.2 years [SD 10.3], p<.001), had a lower proportion of non-Hispanic White participants (84.7% vs 92.4%, p<.001), were less likely to be employed before the pandemic (57.5% vs 86.0%, p<.001), and had a greater comorbidity burden (45.8% vs 64.5% without comorbidity). 613 pwMSRD and 614 controls (63.3% vs. 61.2%; p=.36; **Table 1**) reported a history of acute COVID-19. Compared to controls, pwMSRD were more likely to have had a pre-Omicron infection (44.5% vs 37.0%; p=.008), had a higher rate of severe or critical acute COVID-19 (4.7% vs 0.7%; p<.001), and were more likely to report multiple acute COVID-19 infections (27.1% vs 17.1%; p<.001). Vaccination status was similar between groups (26.1% vs 26.2% not fully vaccinated; p=.25), as was the time from initial infection to survey completion (10.0 months [SD 8.0] vs 9.6 months [SD 8.3], p=.45).

**Table 1.** Participant characteristics. MSRD, multiple sclerosis and related disorder; CCI, Charlson comorbidity index; NMODS, neuromyelitis optica spectrum disorder; MOGAD, myelin oligodendrocyte glycoprotein antibody-associated disease; NID, neuroimmunological disorders; DMT, disease-modifying therapy; RRMS, relapsing remitting multiple sclerosis; PPMS, primary progressive multiple sclerosis; SPMS, secondary progressive multiple sclerosis

| Variables | Variable Category | Overall | MSRD | Control | p-value |
| --- | --- | --- | --- | --- | --- |
| N |  | 1972 | 969 | 1003 |  |
| Age, years; mean (SD) |  | 48.4 (11.7) | 51.8 (12.1) | 45.2 (10.3) | <.001 |
| Sex; n (%) | Female | 1599 (81.1%) | 803 (82.9%) | 796 (79.4%) | .183 |
|  | Male | 370 (18.8%) | 166 (17.1%) | 204 (20.3%) |  |
| Race and ethnicity; n (%) | Non-Hispanic White | 1748 (88.6%) | 821 (84.7%) | 927 (92.4%) | <.001 |
|  | Other | 224 (11.4%) | 148 (15.3%) | 76 (7.6%) |  |
| BMI, kg/m<sup>2</sup>; mean (SD) |  | 33.3 (8.7) | 33.3 (8.7) | 33.2 (8.7) | .753 |
| CCI category; n (%) | None (CCI=0) | 1091 (55.3%) | 444 (45.8%) | 647 (64.5%) | <.001 |
|  | Mild (1≤CCI≤2) | 605 (30.7%) | 305 (31.5%) | 300 (29.9%) |  |
|  | Moderate (3≤CCI≤5) | 235 (11.9%) | 188 (19.4%) | 47 (4.7%) |  |
|  | Severe (CCI≥6) | 41 (2.1%) | 32 (3.3%) | 9 (0.9%) |  |
| Pre-pandemic employment status; n (%) | Employed | 1423 (72.1%) | 560 (57.8%) | 863 (86.0%) | <.001 |
|  | Unemployed | 549 (27.9%) | 409 (42.2%) | 140 (14.0%) |  |
| Neurologic disorder; n (%) | MOGAD | N/A | 2 (0.2%) | N/A | N/A |
|  | MS | N/A | 940 (97.0%) | N/A |  |
|  | NID | N/A | 18 (1.9%) | N/A |  |
|  | NMOSD | N/A | 9 (0.9%) | N/A |  |
| MS subtype | RRMS | N/A | 714 (76.0%) | N/A |  |
|  | SPMS | N/A | 105 (11.1%) | N/A |  |
|  | PPMS | N/A | 65 (6.9%) | N/A |  |
| DMT efficacy; n (%) | None | N/A | 177<br>(18.8%) | N/A | N/A |
|  | Anti-CD20s | N/A | 339<br>(36.1%) | N/A |  |
|  | S1P modulators | N/A | 67 (7.1%) | N/A |  |
|  | Other | N/A | 357<br>(38.0%) | N/A |  |
| Acute COVID-19; n (%) | Present | 1227<br>(62.2%) | 613<br>(63.3%) | 614<br>(61.2%) | .353 |
|  | Absent | 745<br>(37.8%) | 356<br>(36.7%) | 389<br>(38.8%) |  |
| Multiple infection episodes; n (%) |  | 271<br>(22.1%) | 166<br>(27.1%) | 105<br>(17.1%) | <.001 |
| Initial acute COVID-19 detection source; n (%) | PCR | 459<br>(37.4%) | 252<br>(41.1%) | 207<br>(33.7%) | .030 |
|  | Rapid antigen test | 583<br>(47.5%) | 275<br>(44.9%) | 308<br>(50.2%) |  |
|  | Other | 185<br>(15.1%) | 86<br>(14.0%) | 99<br>(16.1%) |  |
| Initial acute COVID-19 severity; n (%) | No symptoms | 45 (3.8%) | 23 (3.6%) | 22 (4.1%) | <.001 |
|  | Mild | 653<br>(53.2%) | 290<br>(47.6%) | 363<br>(58.8%) |  |
|  | Moderate | 495<br>(40.3%) | 271<br>(44.2%) | 224<br>(36.5%) |  |
|  | Severe and critical | 34 (2.7%) | 29 (4.7%) | 5 (0.7%) |  |
| Initial infection wave; n (%) | Before Jan 1, 2022 | 500<br>(40.1%) | 273<br>(44.5%) | 227<br>(37.0%) | .008 |
|  | After Jan 1, 2022 | 727<br>(59.9%) | 340<br>(55.5%) | 387<br>(63.0%) |  |
| Vaccine dose; n (%) | <3 | 513<br>(26.1%) | 253<br>(26.1%) | 263<br>(26.2%) | .245 |
|  | ≥3 | 1456<br>(73.9%) | 716<br>(73.9%) | 740<br>(73.8%) |  |
| Months from the initial infection to survey completion; mean (SD) |  | 9.8 (8.1) | 10.0 (8.0) | 9.6 (8.3) | .450 |

### Post-infection symptoms

The five most frequent new-onset symptoms persisting over 3 months after acute COVID-19 differed between pwMSRD (change or loss of smell, change or loss of taste, shortness of breath, cough with mucus, and breathing difficulty; 6.7-10.4%; **eFigure 1**) and controls (brain fog, fatigue, exercise intolerance, speech and language issues, and memory issues; 5.7-9.9%; **eFigure 2**). After adjusting for covariates and applying Bonferroni correction, pwMSRD had significantly lower adjusted odds of new-onset brain fog (0.3 [0.2, 0.6]), fatigue (0.2 [0.1, 0.3]), and speech and language issues (0.2 [0.1, 0.5]) (**eFigure 3**).

**Figure 3.**
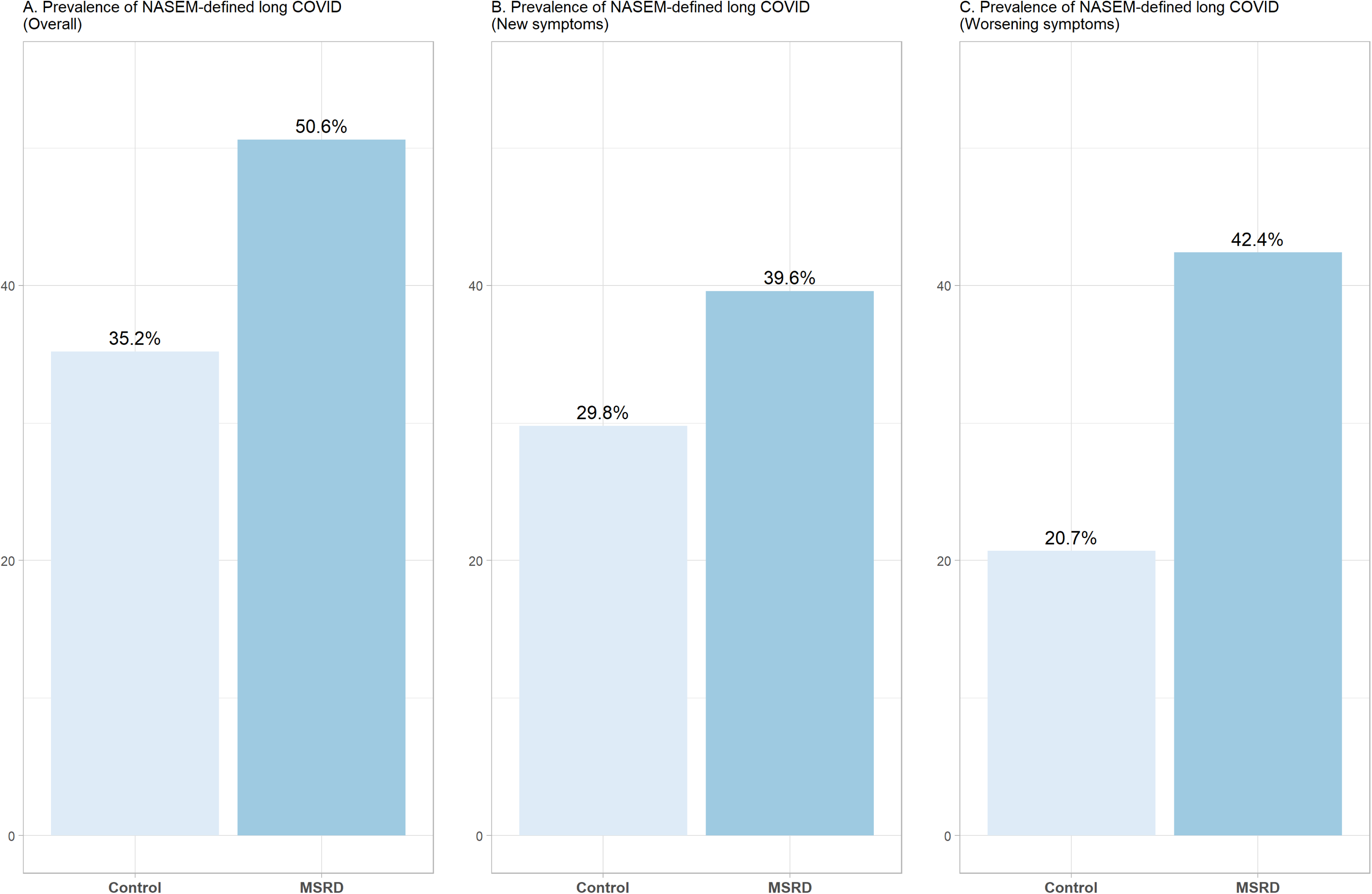
Prevalence of NASEM-defined long COVID.

The most frequent worsening symptoms (>10% prevalence) persisting over 3 months after acute COVID-19 in pwMSRD were fatigue, brain fog, weakness, difficulty concentrating, memory issues, dizziness, speech and language issues, joint pain, muscle aches, muscle spasms, change in sensation, exercise intolerance, and insomnia (**eFigure 4**), none of which exceeded 10% prevalence in controls (**eFigure 5**). PwMSRD had significantly higher adjusted odds of worsening symptoms **(eFigure 6**), including dizziness (aOR=15.7), weakness (aOR=14.9), muscle spasms (aOR=9.3), change in sensation (aOR=8.0), speech and language issues (aOR=7.0), difficulty concentrating (aOR=5.3), brain fog (aOR=6.0), fatigue (aOR=4.9), bladder control issues (aOR=4.2), muscle aches (aOR=3.4), and joint pain (aOR=2.8).

**Figure 4.**
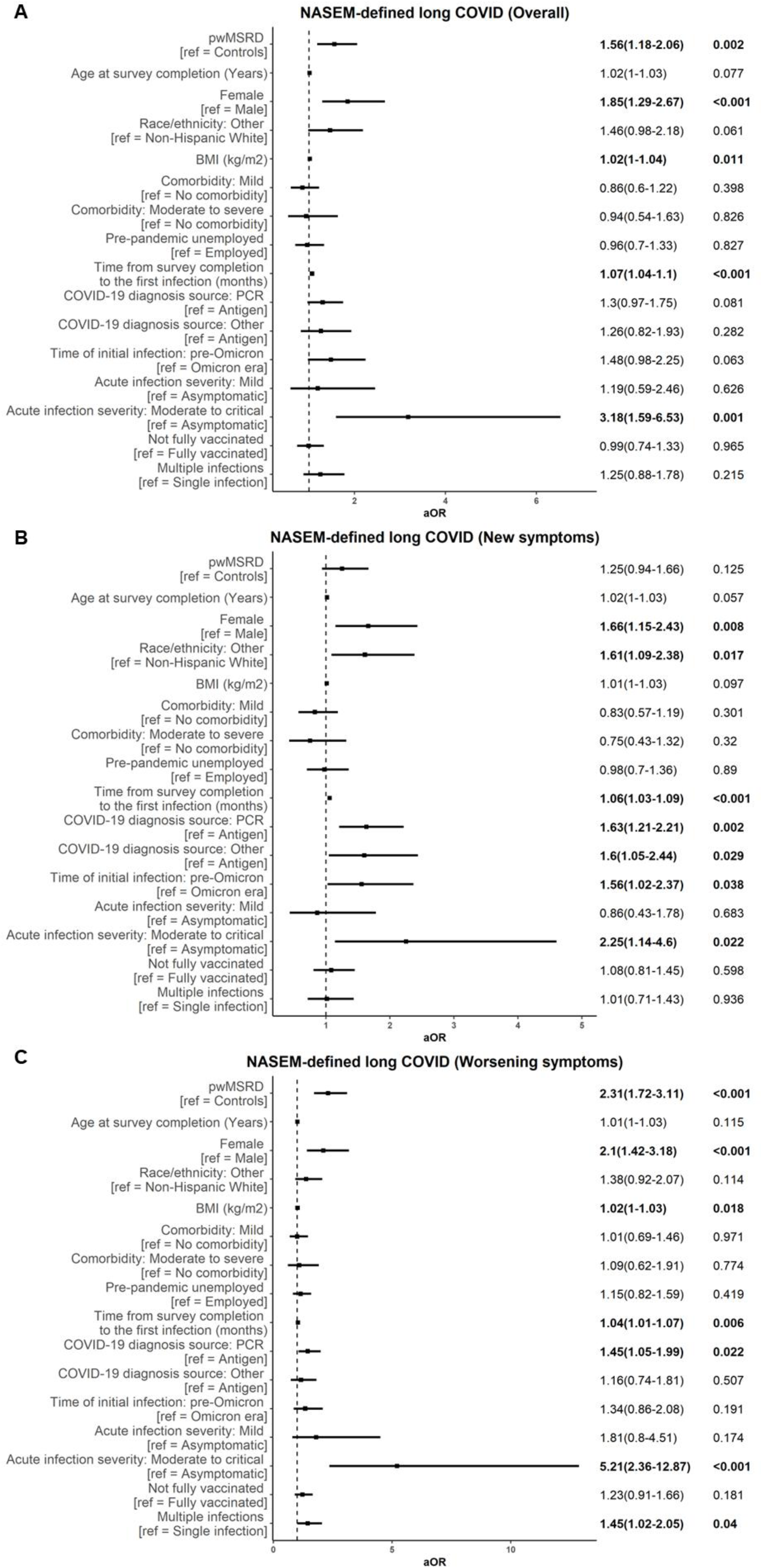
Associations between factors and NASEM-defined long COVID. Adjusted odds ratios (aORs) with 95% confidence intervals (CIs) from multivariable logistic regression models evaluating factors associated with (A) overall NASEM-defined long COVID, (B) long COVID based on new symptoms, and (C) long COVID based on worsening symptoms.

**Figure 5.**
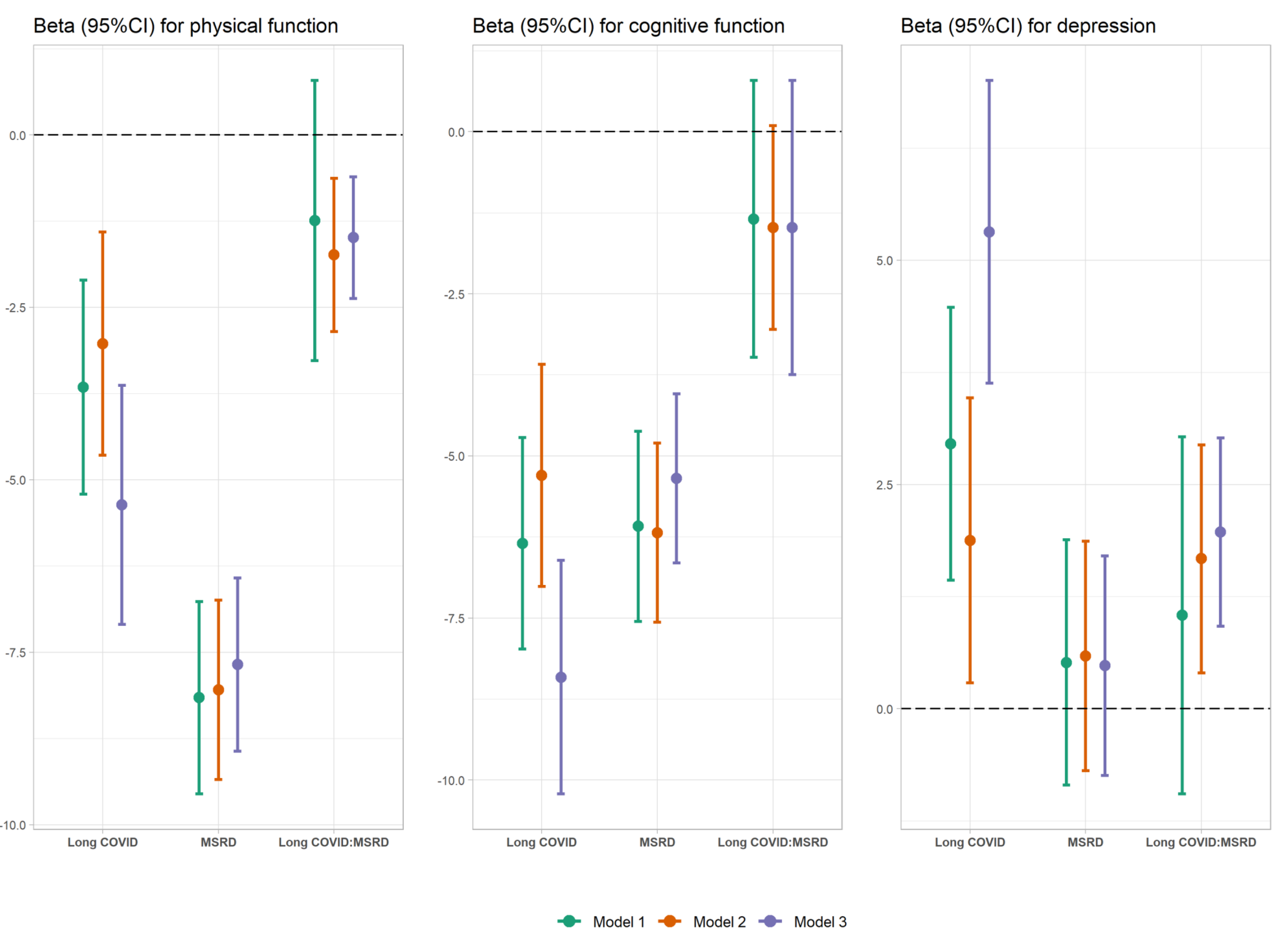
Multivariable linear regression for PROMIS physical function (left), cognitive function (middle), and depression (right). Model 1, NASEM-defined long COVID overall as the main exposure; Model 2, NASEM-defined long COVID on new symptoms as the main exposure; Model 3, NASEM-defined long COVID on worsening symptoms as the main exposure.

### Associations between MSRD status and long COVID

Using the NASEM definition of long COVID overall, 310 (50.6%) pwMSRD met the criteria compared to 216 (35.2%) controls (p<.001; **Figure 3A**). When based on new symptoms, the long COVID prevalence was 39.6% in pwMSRD vs 29.8% in controls (p<.001; **Figure 3B**). When based on worsening symptoms, the long COVID prevalence was 42.4% in pwMSRD vs 20.7% in controls (p<.001; **Figure 3C**). After adjusting for confounders, pwMSRD had higher odds of developing long COVID overall (aOR=1.6 [1.2, 2.1]; **Figure 4A**) and long COVID based on worsening symptoms (aOR=2.3 [1.7, 3.1]; **Figure 4C**]), whereas the association with long COVID based on new symptoms was attenuated and no longer statistically significant (aOR=1.3 [0.9, 1.7]; **Figure 4B**). Female and moderate-to-critical acute COVID-19 severity (vs asymptomatic infection) were consistently associated with higher odds of long COVID across all three definitions (**Figure 4A-C**).

### Associations between long COVID and PROMIS T-scores

1,115 (90.9%) participants who completed all three PROMIS assessments were included in the PRO analysis (**Figure 2**). Compared to the excluded participants, the participants included were more likely to be employed and develop long COVID, while other demographics were similar (**eTable 3**). Multivariable linear regression showed that long COVID (overall, new, or worsening) was associated with significantly decreased physical function (3.0-5.4 points) and cognitive function (6.2-8.4 points) and with significantly increased depression severity (1.9-5.3 points) (**Figure 5; eTable 4**). Notably, MSRD status modified the association of long COVID with PROs: pwMSRD with long COVID experienced greater physical function declines and depression severity than controls (interaction p<.05; **eTable 4**).

Subgroup results: Long COVID and disability in pwMS

Among pwMS, 282 (50.3%) had NASEM-defined long COVID overall, 220 (39.2%) based on new symptoms, and 236 (42.1%) based on worsening symptoms (**Table 2**). Female, worse pre-COVID disability, and worse acute infection severity were significant contributors to long COVID development (all LRT p-values<.05), while MS subtype and DMT use were not. Analyses restricted to new or worsening symptoms yielded similar findings regarding key contributing factors (**Table 2**). Patients with long COVID exhibited greater patient-reported disability accumulation based on PDDS at the time of the survey: aRR=1.2 [1.2, 1.5] for long COVID overall; aRR=1.2 [1.1, 1.4] for long COVID based on new symptoms; and aRR=1.2 [1.1, 1.4] for long COVID based on worsening symptoms.

**Table 2.** Contributing factors to long COVID in multiple sclerosis (MS) patients. ^a^ P-values of likelihood ratio tests (LRT) comparing full and reduced models RRMS, relapsing-remitting multiple sclerosis; PMS, progressive multiple sclerosis (primary and secondary); S1P modulators, sphingosine-1-phosphate receptor modulator; PDDS, patient determined disease steps

| Variables | Variable Categories | NASEM-defined long COVID (Overall)<br>n = 282 (50.3%) |  | NASEM-defined long COVID (New symptoms)<br>n = 220 (39.2%) |  | NASEM-defined long COVID (Worsening symptoms)<br>n = 236 (42.1%) |  |
| --- | --- | --- | --- | --- | --- | --- | --- |
|  |  | OR (95%CI) | p-value<sup>a</sup> | OR (95%CI) | p-value<sup>a</sup> | OR (95%CI) | p-value<sup>a</sup> |
| Age at survey completion (Years) | - | 1(0.97-1.02) | 0.96 | 1.01(0.98-1.04) | 0.549 | 0.99(0.95-1.02) | 0.524 |
| Sex [ref = Male] | Female | **2.96(1.58-5.82)** | **<0.001** | **2.01(1.01-4.49)** | **0.044** | **2.54(1.02-7.45)** | **0.044** |
| Race/ethnicity [ref = Non-Hispanic White] | Other | **1.72(1.03-2.9)** | **0.038** | **1.77(1.02-3.03)** | **0.041** | 1.41(0.72-2.67) | 0.305 |
| Body mass index (kg/m2) | - | 1.01(0.99-1.04) | 0.313 | 1(0.97-1.03) | 0.974 | 1.02(0.98-1.05) | 0.327 |
| Comorbidity [ref = No comorbidity] | Mild | 1.49(0.9-2.49) | 0.295 | 1.19(0.69-2.06) | 0.067 | 1.72(0.86-3.48) | 0.274 |
|  | Moderate to severe | 1.24(0.59-2.64) |  | 0.5(0.21-1.17) |  | 1.91(0.68-5.62) |  |
| Pre-pandemic employment [ref = Employed] | Unemployed | 0.9(0.57-1.41) | 0.652 | 1.14(0.69-1.89) | 0.601 | 0.93(0.5-1.68) | 0.801 |
| MS subtype [ref = RRMS] | PMS | 0.8(0.4-1.59) | 0.52 | 0.72(0.32-1.58) | 0.419 | 0.87(0.35-2.09) | 0.75 |
| DMT use [ref = None] | Anti-CD20s | 0.62(0.34-1.12) | 0.3 | 0.81(0.42-1.61) | 0.807 | 0.57(0.27-1.2) | 0.344 |
|  | S1P modulators | 0.61(0.25-1.42) |  | 0.72(0.25-1.95) |  | 0.55(0.14-1.81) |  |
|  | Other | 0.57(0.32-1.03) |  | 1(0.52-1.93) |  | 0.5(0.24-1.07) |  |
| Baseline PDDS scale | - | **1.17(1.04-1.32)** | **0.008** | **1.3 (1.11-1.54)** | **0.039** | **1.19(1.02-1.38)** | **0.026** |
| Time from survey completion to the first | - | 0.99(0.95-1.03) | 0.599 | 1.02(0.98-1.07) | 0.391 | 1.04(0.99-1.1) | 0.121 |
| infection (months) |  |  |  |  |  |  |  |
| COVID-19 detection source<br>[ref = Antigen] | PCR | 1.47(0.93-2.32) | 0.251 | 1.17(0.7-1.95) | 0.783 | 1.13(0.62-2.08) | 0.209 |
|  | Other | 1.29(0.65-2.54) |  | 0.97(0.44-2.04) |  | 0.51(0.18-1.31) |  |
| Time of initial infection<br>[ref = Omicron era] | pre-Omicron | 1.4(0.74-2.62) | 0.299 | 1.25(0.61-2.53) | 0.543 | 0.53(0.21-1.27) | 0.158 |
| Acute infection severity<br>[ref = Asymptomatic] | Mild | **1.52(0.82-2.82)** | <0.001 | **1.55(0.58-4.14)** | <0.001 | **1.14(0.11-11.82)** | <0.001 |
|  | Moderate to critical | **5.09(3.02-8.58)** |  | **4.1(2.04-8.24)** |  | **7.32(3.16-16.96)** |  |
| Vaccination<br>[ref = Fully vaccinated] | Not fully vaccinated | 1.24(0.8-1.93) | 0.334 | 1.19(0.73-1.93) | 0.481 | 1.65(0.93-2.91) | 0.088 |
| Multiple infections<br>[ref = No] | Yes | 1.28(0.78-2.1) | 0.319 | 0.86(0.49-1.49) | 0.596 | 1.5(0.79-2.84) | 0.212 |

### Sensitivity results: RECOVER-defined Long COVID

When using the RECOVER criteria, the long COVID prevalence declined in both pwMSRD (7.7-17.5%) and controls (1.8-8.0%) (**eFigure 7**). The increased odds of long COVID in pwMSRD vs controls as driven by worsening symptoms remained consistent (aOR = 3.5 [1.7, 7.5]; **eFigure 8**). Similar to NASEM-defined long COVID, RECOVER-defined long COVID was also associated with worse PROs (**eFigure 9; eTable 5**) and a 15.3%-16.7% greater disability accumulation in pwMS.

## Discussion

In this multicenter study of pwMSRD and controls without neuroinflammatory disorders, we systematically surveyed long COVID-19 symptoms. We directly compared each symptom and long COVID development (as defined by the 2024 NASEM and the 2023 RECOVER criteria) between pwMSRD and controls. We refined the NASEM and RECOVER criteria by separately analyzing and distinguishing long COVID based on new-onset and worsening symptoms, which was applicable for chronic neurological disorders such as MSRD. PwMSRD exhibited a higher prevalence of worsening systemic, musculoskeletal, and neurological symptoms from their pre-COVID baseline compared to controls. After adjusting for potential confounders, pwMSRD had 56% higher odds of long COVID overall than controls, with a stronger association (OR=2.3) when long COVID was based on worsening symptoms and a weaker association (OR=1.3) when based on new symptoms.

Research on long COVID susceptibility and its clinical manifestations in MSRD has been limited, and few distinguished newly onset symptoms and worsening symptoms attributable to long COVID. The main finding of increased odds of long COVID in pwMSRD primarily driven by worsening symptoms aligned with a previous study reporting that pwMS were more likely to experience post-COVID weakness, mobility difficulties, and cognitive dysfunction than controls, after accounting for pre-existing symptoms.^21^ Acute or chronic systemic infections can trigger symptomatic exacerbations in MSRD.^22^ Potential mechanisms of exacerbation include systemic inflammatory response across organs (including the central nervous system) in the setting of superantigens as well as molecular mimicry, epitope spreading, bystander activation in the presence of cryptic antigens.^23^

Assessing long COVID in pwMSRD was challenging due to multiple factors, which we addressed to enhance the study rigor. First, a standardized long COVID definition applicable to pwMSRD is lacking, given the potential overlap of symptoms between MSRD and long COVID.^2,24^ To mitigate misclassification bias, we performed primary analyses using the latest NASEM criteria and performed sensitivity analyses using the widely used RECOVER criteria. The study findings were largely comparable across the two criteria. By distinguishing newly onset symptoms from worsening pre-existing symptoms, we aimed to clarify the clinical presentation of long COVID in pwMSRD versus controls. Second, the NASEM and RECOVER criteria differed. As expected, NASEM-defined long COVID prevalence was higher than RECOVER-defined long COVID prevalence due to fewer restrictions on the type and number of symptoms required. In this study, 17.5% of pwMSRD developed RECOVER-defined long COVID, consistent with the 15.9% reported by Salter et al that also used the RECOVER scoring system.^10^ Future research is needed to rigorously standardize and optimize long COVID definitions specific to the MSRD populations.^13^ Third, pwMSRD differed from controls in demographic and clinical profile, acute COVID-19 severity, and vaccination status, all of which were potential confounders.^25,26^ We systematically compared these factors between groups and the analyses adjusted for them to reduce confounding bias. However, residual confounding could remain, and we did *not* conclude a causal relationship between MSRD and long COVID, which warrants future investigation.

The PRO analysis in this study was consistent with the inverse association between long COVID and functional capacity in the general population. More importantly, we found a synergism between MSRD and long COVID on physical function and depression, underscoring the need for better management of long COVID in pwMSRD. While prior research suggested that acute COVID-19 infection does not immediately affect disability, our findings indicated that long COVID contributed to accelerated long-term disability worsening or accumulation.^27^ Future longitudinal studies with extended follow-up are necessary to disentangle the relationships between acute infection, immediate disability changes, long COVID, and long-term disability progression. Interestingly, progressive MS and BCD treatment, which were associated with severe acute COVID-19, did not significantly contribute to long COVID development in this study. This could be due to the stronger impact of acute infection severity, which was accounted by our analyses.^2^

This study has several strengths. First, the multicenter design provided a large, representative sample of pwMSRD and controls from the Northeastern and Mid-Atlantic U.S. Second, the inclusion of a control group enabled direct comparisons, aiding future research and clinical guidance. Third, the post-infection symptom survey systematically assessed a broad range of symptoms while distinguishing between new and worsening symptoms, facilitating a nuanced comparison of symptoms manifestations. Finally, we made efforts to reduce misclassification bias and enhance robustness of findings by assessing different long COVID definitions.

There are also limitations. The retrospective survey design might introduce selection and recall biases. Second, the cross-sectional study required cautious interpretation as we did not establish causality. Finally, the study findings may not generalize to people who did not volunteer to answer this survey and racial and ethnic minorities, highlighting the need for future validation studies.

In summary, pwMSRD exhibited heightened odds for long COVID, particularly due to worsening pre-existing symptoms, which exacerbated functional decline and accelerated disability progression. Our study provided real-world evidence of long COVID development and consequences in the vulnerable population of MSRD and highlighted the importance of careful clinical evaluation and management of long COVID in the post-pandemic era.

## Data Availability

De-identified data are available upon request to the corresponding author and with permission from the participating institutions.

## Acknowledgment

We thank all research participants for their efforts.

## Author contributions

C.H., J.S., L.M., and Z.X. contributed to the conception and design of the study; A.B., C.P., C.R., B.G., P.J., and E.L. contributed to the acquisition and analysis of data; M.D., S.B., K.K., K.O., E.S., and E.W. contributed to drafting the text or preparing the figures; C.H., J.S., L.M., and Z.X. contributed to drafting and editing of the manuscript.

## Data sharing statement

Individual de-identified participant data and data dictionaries will be shared upon request and with permission from the participating institutions. Requests for data access should be directed to the corresponding author, and agreements will be facilitated to ensure the ethical use of the shared data. Access to the data will be granted to qualified researchers whose proposals have been reviewed and approved by the corresponding author and participating institutions. Approved data will be provided via a secure transfer mechanism, ensuring compliance with all applicable ethical and institutional guidelines. Code for analyses and figures is publicly available at: https://github.com/xialab2016/Post-COVID-Sequelae.

