## Supplemental Materials for "Long COVID in people with multiple sclerosis and related disorders: a multicenter cross-sectional study"

### Supplementary Tables

**eTable 1. Post-infection symptoms surveyed and their classification within the RECOVER scoring system**

| Organ system | Symptoms | Classification into RECOVER scoring components |
|---|---|---|
| Systemic symptoms | Chills | |
| | Fatigue | Fatigue |
| | Elevated temperatures | |
| | Fever | |
| | Low temperature | |
| | Exercise intolerance | Post-exertional malaise |
| Respiratory symptoms | Breathing difficulties | |
| | Requiring supplemental oxygen | |
| | Cough with mucus | Chronic cough |
| | Cough with blood | Chronic cough |
| | Dry cough | Chronic cough |
| | Shortness of breath | |
| | Wheezing | |
| | Sneezing | |
| Gastrointestinal symptoms | Abdominal pain | Gastrointestinal |
| | Constipation | Gastrointestinal |
| | Diarrhea | Gastrointestinal |
| | Loss of appetite | Gastrointestinal |
| | Reflux | Gastrointestinal |
| | Nausea | Gastrointestinal |
| | Vomiting | Gastrointestinal |

| | | |
|---|---|---|
| Reproductive / Genitourinary symptoms | Menstrual changes | |
| | Bladder control issues | |
| Cardiovascular symptoms | Bradycardia | |
| | Tachycardia | |
| | Fainting | |
| | Palpitations | Palpitations |
| | Chest pain | Chest pain |
| Dermatologic symptoms | Change in color of hands and/or feet | |
| | Dermatographia | |
| | Skin rashes | |
| | Peeling skin | |
| | Petechiae | |
| | Alopecia | Hair loss |
| | Change in nails | |
| | Other allergy skin issues | |
| Musculoskeletal symptoms | Bone aches | |
| | Joint pain | |
| | Muscle aches | |
| | Muscle spasms | |
| | Tightness of chest | |
| Immunologic symptoms | Allergies | |
| | Anaphylaxis | |
| HEENT symptoms | Hearing loss | |
| | Tinnitus | |
| | Runny nose | |

| | | |
|---|---|---|
| | Thirst | Thirst |
| | Vision symptoms | |
| | Change or loss of smell | Smell/taste |
| | Change or loss of taste | Smell/taste |
| | Nasal congestion | |
| Neuropsychiatric cognitive symptoms | Acute confusion | |
| | Brain fog | Brain fog |
| | Memory issues | |
| | Slurring words | |
| | Speech and language issues | |
| | Difficulty concentrating | |
| | Getting lost in familiar place | |
| Neuropsychiatric sensorimotor symptoms | Change in sensation | |
| | Dizziness | Dizziness |
| | Neuralgia | Abnormal movements |
| | Tremors | Abnormal movements |
| | Vibration sensation | Abnormal movements |
| | Weakness | |
| Neuropsychiatric other symptoms | Anxiety | |
| | Depression | |
| | Sexual desire or capacity | Sexual desire or capacity |
| | Insomnia | |
| | Hypersomnia | |
| | Sleep apnea | |

| | |
|---|---|
| | Hallucinations |

**eTable 2. Participant characteristics of survey responder vs non-consenter or non-responder**

| | Total | Non-consenter OR Non-responder | Responder |
|---|---|---|---|
| N | 3,527 | 1,555 | 1,972 |
| Year of birth; Median | 1972 | 1973 | 1969 |
| pwMSRD; n (%) | 2,190 | 1,221 (78.5%) | 969 (49.1%) |
| Female; n (%) | 2,518 | 919 (59.1%) | 1,599 (81.1%) |
| Non-Hispanic White; n (%) | 3,001 | 1,253 (80.6%) | 1,748 (88.6%) |

**eTable 3. Characteristics between participants included vs excluded for analysis of patient-reported outcomes**

| | Include (n = 1,115) | Exclude (n = 112) | p-value |
|---|---|---|---|
| Age at survey, years; Mean (SD) | 46.5 (12.5) | 47. 0 (11.1) | 0.705 |
| Female; n (%) | 911 (81.7%) | 99 (88.4%) | 0.101 |
| Non-Hispanic White; n (%) | 983 (88.2%) | 93 (83.0%) | 0.155 |
| BMI; Mean (SD) | 32.9 (8.7%) | 33.3 (8.6%) | 0.622 |
| No comorbidity; n (%) | 661 (59.3%) | 70 (62.5%) | 0.490 |
| Employed; n (%) | 875 (78.5%) | 41 (36.6%) | < 0.001 |
| Long COVID; n (%) | 304 (27.3%) | 19 (17.0%) | 0.025 |

**eTable 4. Estimates (95%CI) of multivariable linear models between NASEM-defined long COVID status and PROMIS outcomes**

| | Beta (95% CI) | p-value |
|---|---|---|
| | Physical function | |
| MSRD | -8.16 (-9.55, -6.77) | < 0.001 |
| Long COVID (Overall) | -3.66 (-5.21, -2.11) | < 0.001 |
| MSRD: Long COVID (Overall) | -1.24 (-3.27, 0.79) | 0.102 |
| MSRD | -8.04 (-9.35, -6.74) | < 0.001 |
| Long COVID (New symptoms) | -3.03 (-4.64, -1.41) | < 0.001 |
| MSRD: Long COVID (New symptoms) | -1.74 (-2.85, -0.63) | 0.037 |
| MSRD | -7.68 (-8.94, -6.42) | < 0.001 |
| Long COVID (Worsening symptoms) | -5.37 (-7.1, -3.63) | < 0.001 |
| MSRD: Long COVID (Worsening symptoms) | -1.49 (-2.37, -0.61) | 0.040 |
| | Cognition | |
| MSRD | -6.09 (-7.55, -4.62) | < 0.001 |
| Long COVID (Overall) | -6.35 (-7.98, -4.72) | < 0.001 |
| MSRD: Long COVID (Overall) | -1.35 (-3.48, 0.79) | 0.873 |
| MSRD | -6.18 (-7.57, -4.8) | < 0.001 |
| Long COVID (New symptoms) | -5.3 (-7.02, -3.59) | < 0.001 |
| MSRD: Long COVID (New symptoms) | -1.04 (-3.27, 1.19) | 0.763 |
| MSRD | -5.35 (-6.65, -4.04) | < 0.001 |
| Long COVID (Worsening symptoms) | -8.41 (-10.21, -6.61) | 0.001 |
| MSRD: Long COVID (Worsening symptoms) | -1.48 (-3.05, 0.09) | 0.169 |
| | Depression | |
| MSRD | 0.52 (-0.85, 1.88) | 0.46 |
| Long COVID (Overall) | 2.95 (1.43, 4.47) | < 0.001 |
| MSRD: Long COVID (Overall) | 1.04 (-0.95, 3.03) | 0.305 |
| MSRD | 0.59 (-0.69, 1.87) | 0.368 |

| | | |
|---|---|---|
| Long COVID (New symptoms) | 1.88 (0.29, 3.46) | 0.021 |
| MSRD: Long COVID (New symptoms) | 1.67 (0.4, 2.94) | 0.021 |
| MSRD | 0.48 (-0.74, 1.7) | 0.441 |
| Long COVID (Worsening symptoms) | 5.32 (3.63, 7) | < 0.001 |
| MSRD: Long COVID (Worsening symptoms) | 1.97 (0.92, 3.02) | 0.014 |

**eTable 5. Estimates (95%CI) of multivariable linear models between RECOVER-defined long COVID status and PROMIS outcomes**

| | Beta (95% CI) | p-value |
|---|---|---|
| | Physical function | |
| MSRD | -7.97 (-9.17, -6.78) | < 0.001 |
| Long COVID (Overall) | -5.41 (-7.28, -3.55) | < 0.001 |
| MSRD: Long COVID (Overall) | -1.92 (-4.38, 0.53) | 0.102 |
| MSRD | -8.38 (-9.53, -7.24) | < 0.001 |
| Long COVID (New symptoms) | -4.6 (-6.74, -2.47) | < 0.001 |
| MSRD: Long COVID (New symptoms) | -3.01 (-5.75, -0.27) | 0.031 |
| MSRD | -7.83 (-8.92, -6.74) | < 0.001 |
| Long COVID (Worsening symptoms) | -8.42 (-12.38, -4.46) | < 0.001 |
| MSRD: Long COVID (Worsening symptoms) | -4.43 (-8.82, -0.05) | 0.048 |
| | Cognition | |
| MSRD | -5.33 (-6.61, -4.05) | < 0.001 |
| Long COVID (Overall) | -6.43 (-8.43, -4.42) | < 0.001 |
| MSRD: Long COVID (Overall) | -0.2 (-2.26, 1.86) | 0.873 |
| MSRD | -5.93 (-7.16, -4.69) | < 0.001 |
| Long COVID (New symptoms) | -5.15 (-7.47, -2.83) | < 0.001 |
| MSRD: Long COVID (New symptoms) | -0.46 (-2.51, 1.6) | 0.763 |
| MSRD | -5.44 (-6.62, -4.27) | < 0.001 |
| Long COVID (Worsening symptoms) | -6.97 (-11.21, -2.72) | 0.001 |
| MSRD: Long COVID (Worsening symptoms) | -0.02 (-4.72, 4.68) | 0.993 |
| | Depression | |
| MSRD | 0.71 (-0.47, 1.89) | 0.239 |
| Long COVID (Overall) | 4.54 (2.7, 6.39) | < 0.001 |
| MSRD: Long COVID (Overall) | 0.24 (-2.04, 2.53) | 0.835 |
| MSRD | 1.07 (-0.07, 2.2) | 0.065 |

| | | |
|---|---|---|
| Long COVID (New symptoms) | 2.82 (0.69, 4.95) | 0.010 |
| MSRD: Long COVID (New symptoms) | 0.95 (-1.78, 3.68) | 0.493 |
| MSRD | 0.8 (-0.27, 1.87) | 0.145 |
| Long COVID (Worsening symptoms) | 5.93 (2.05, 9.81) | 0.003 |
| MSRD: Long COVID (Worsening symptoms) | 0.35 (-3.95, 4.65) | 0.873 |

### Supplementary Figures

**eFigure 1. Bar diagram of prevalence of each post-infection new-onset symptom in pwMSRD.**

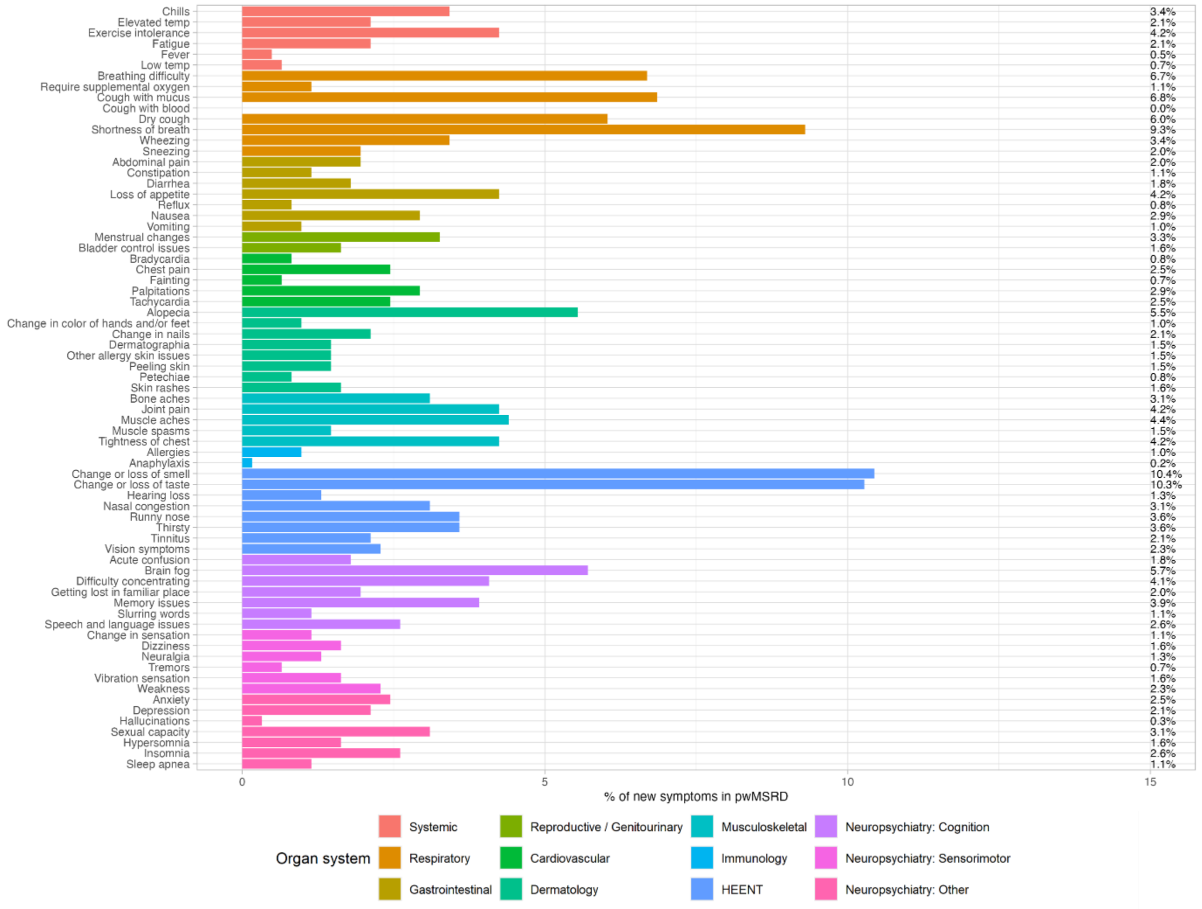

**eFigure 2. Bar diagram of prevalence of each post-infection new-onset symptom in controls.**

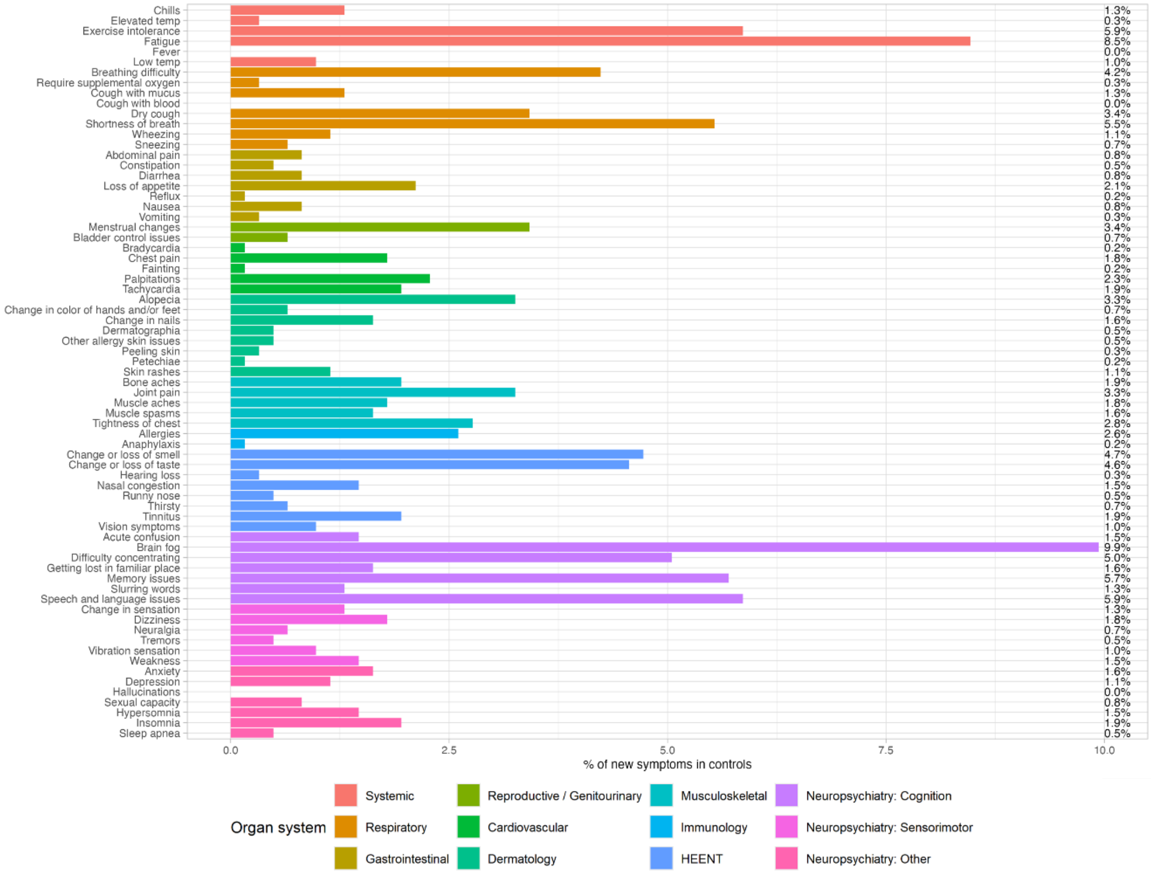

**eFigure 3. Estimated aORs and 95% CIs (gray bars) from multivariable logistic regression comparing adjusted odds of new symptoms between pwMSRD and controls. Symptoms in red reached statistical significance after Bonferroni correction.**

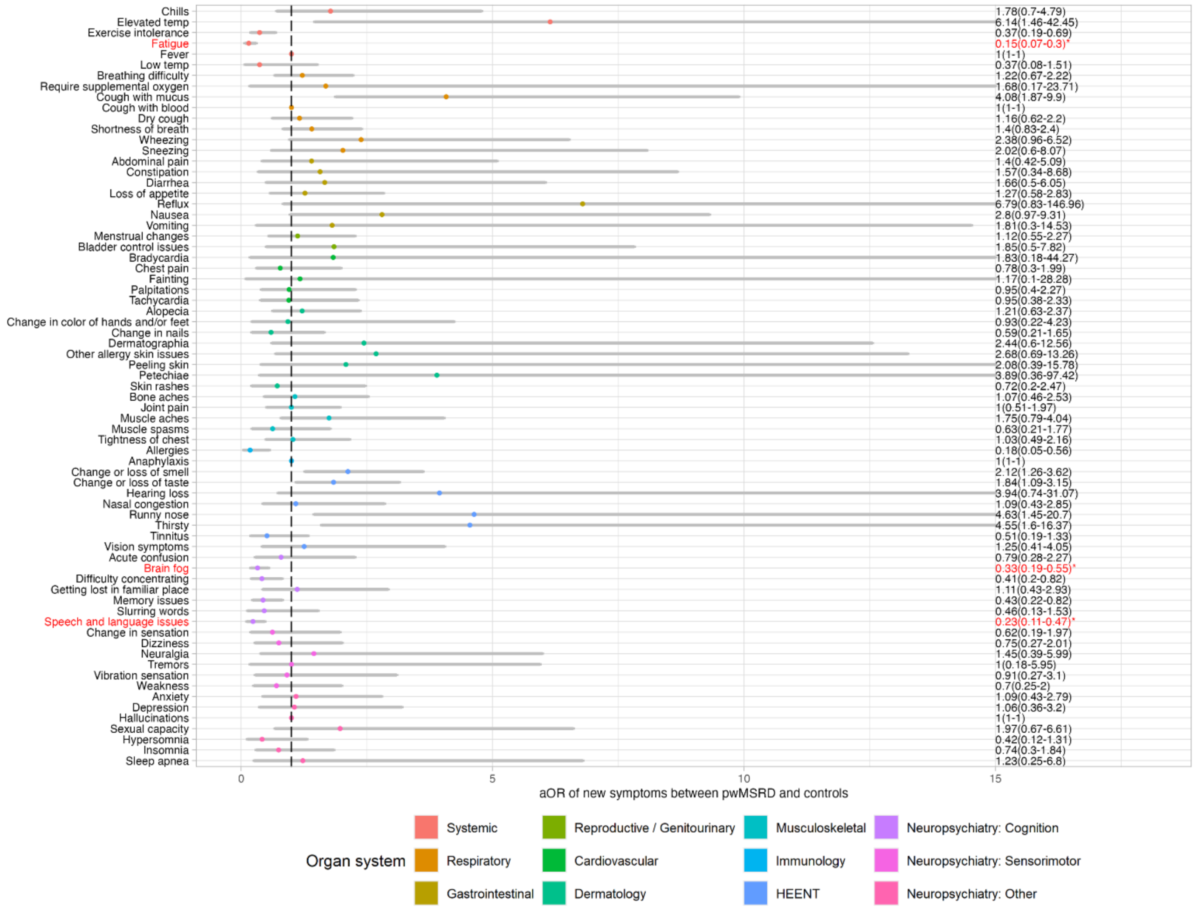

**eFigure 4. Bar diagram of prevalence of each post-infection worsening symptom from pre-COVID in pwMSRD.**

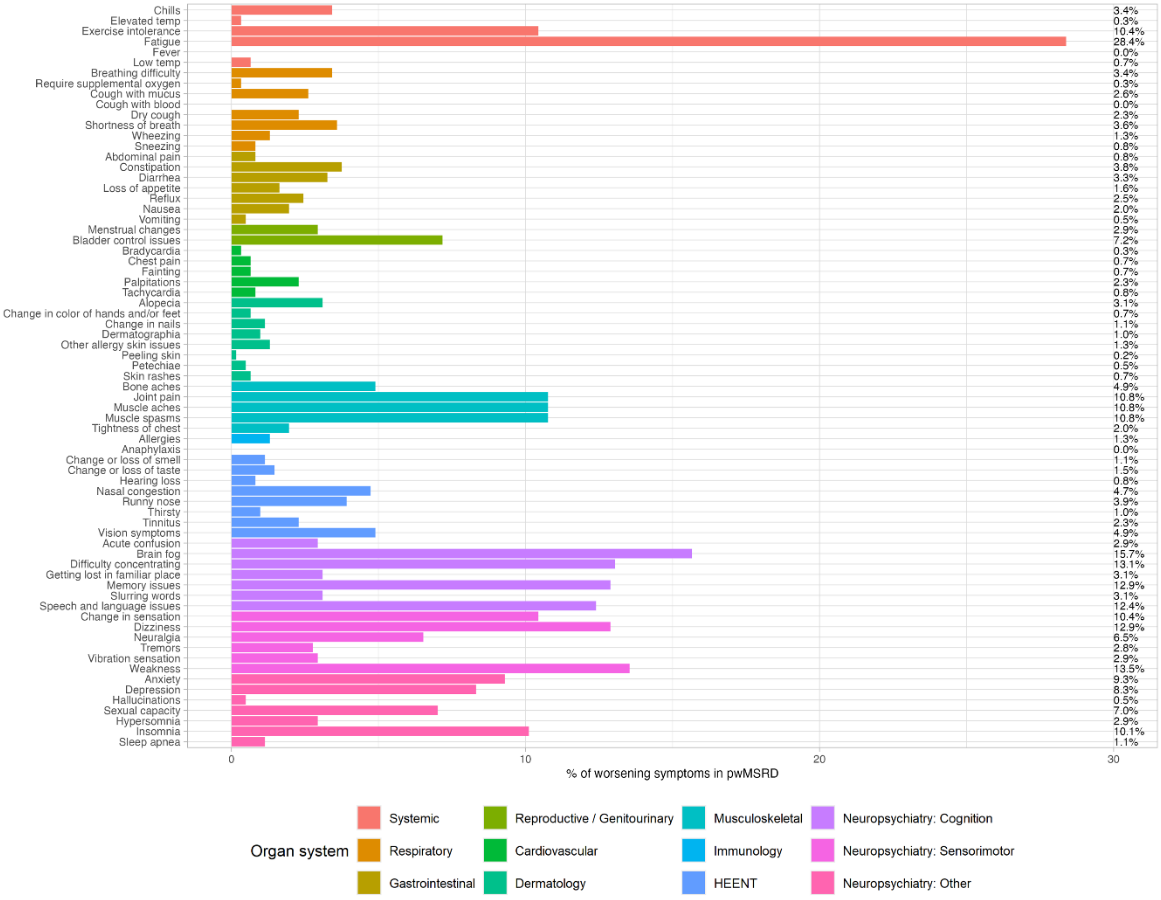

**eFigure 5. Bar diagram of prevalence of each post-infection worsening symptom from pre-COVID in controls.**

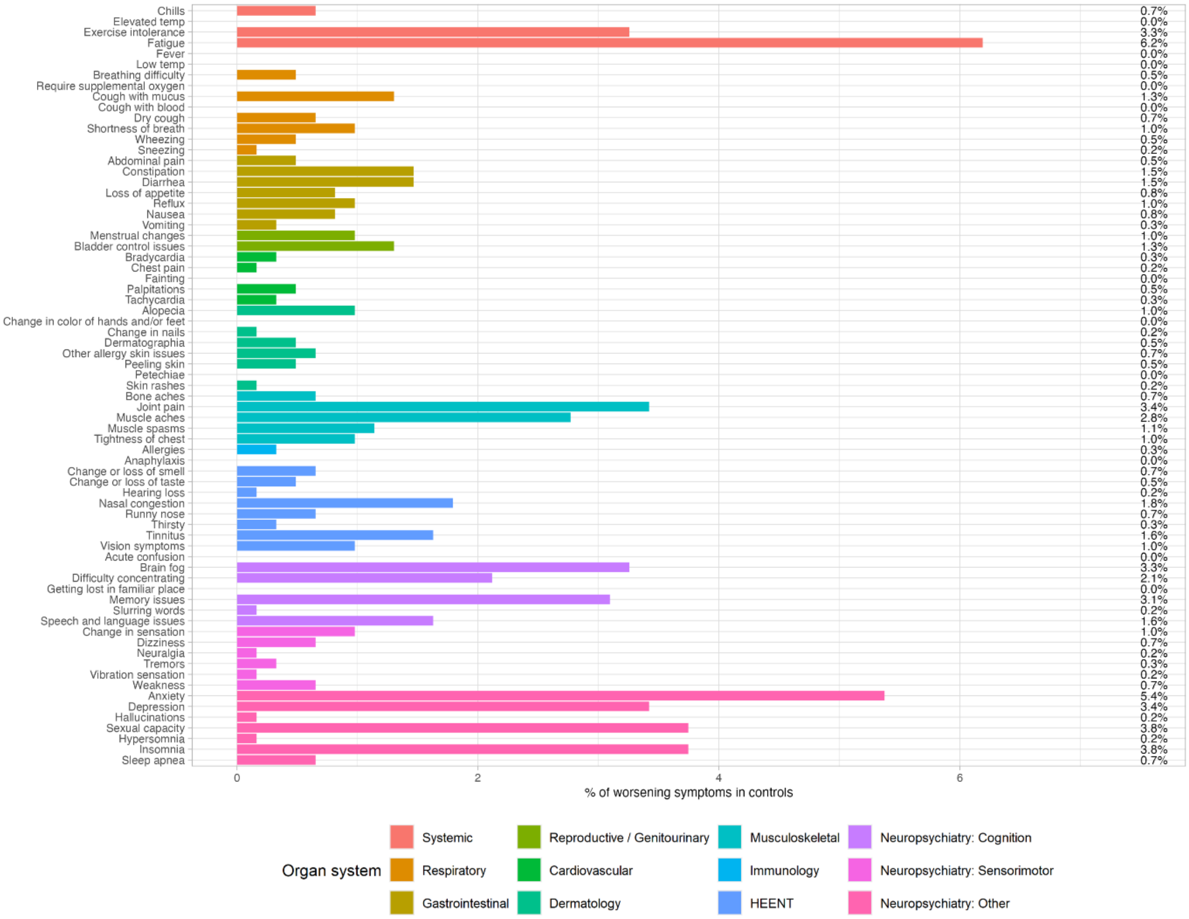

**eFigure 6. Estimated aORs and 95% CIs (gray bars) from multivariable logistic regression comparing the adjusted odds of worsening symptoms between pwMSRD and controls. Symptoms in red reached statistical significance after Bonferroni correction.**

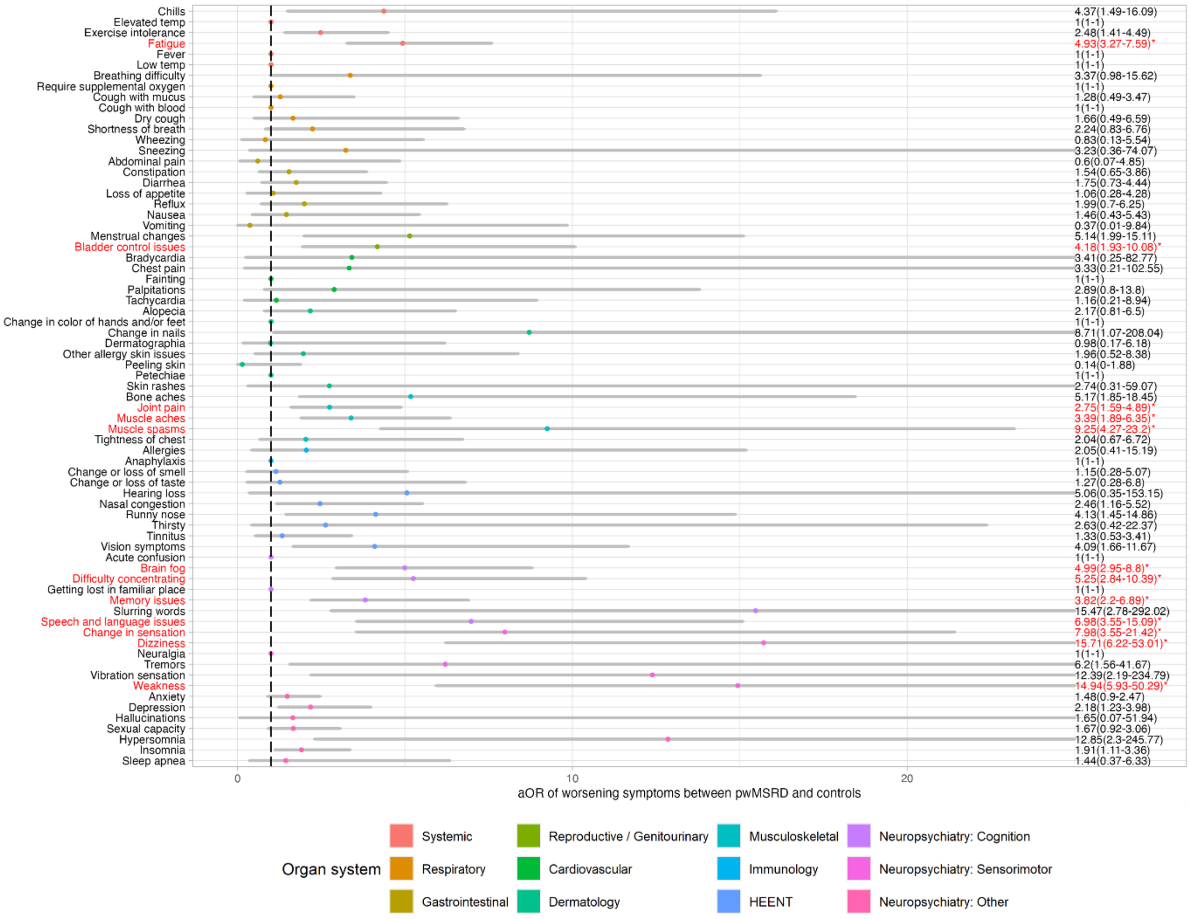

### eFigure 7. Prevalence of RECOVER-defined long COVID.

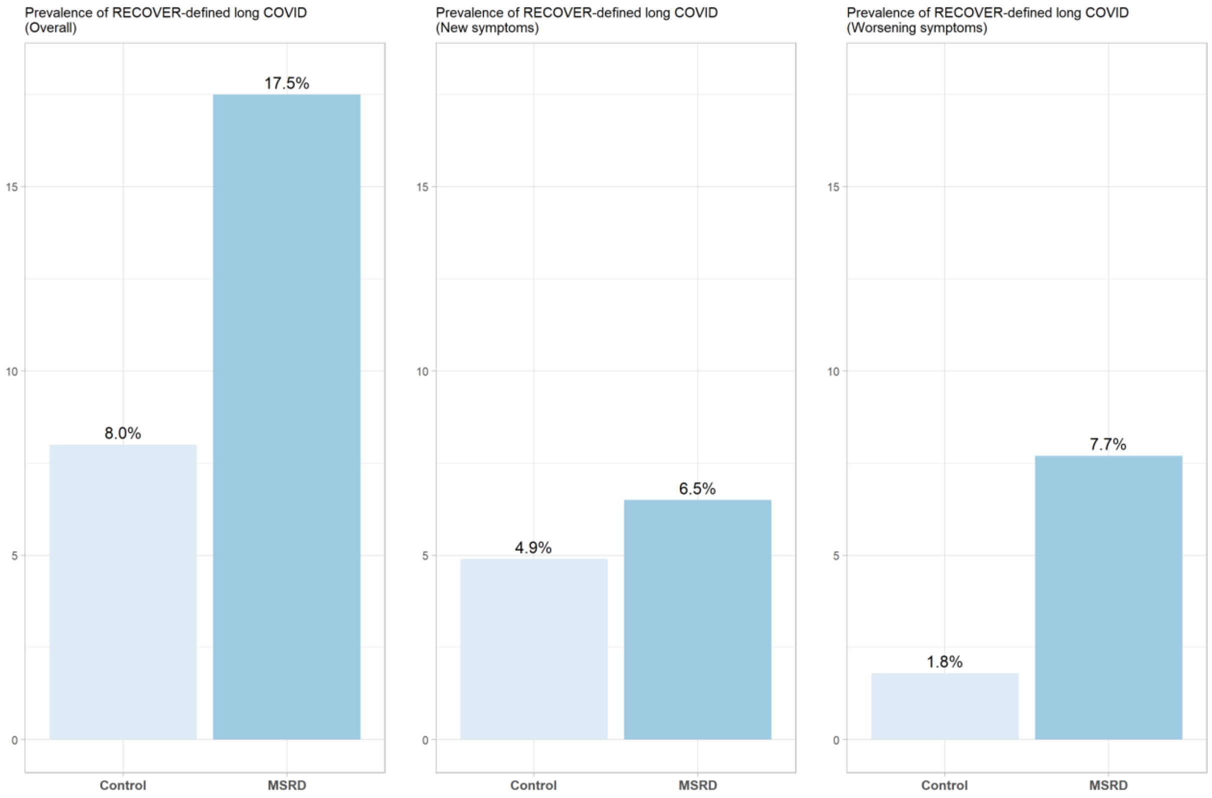

eFigure 8. Results (aOR [95% CI] and p-value) of multivariable logistic regression for (A) overall long COVID, (B) long COVID based on new symptoms, and (C) long COVID based on worsening symptoms.

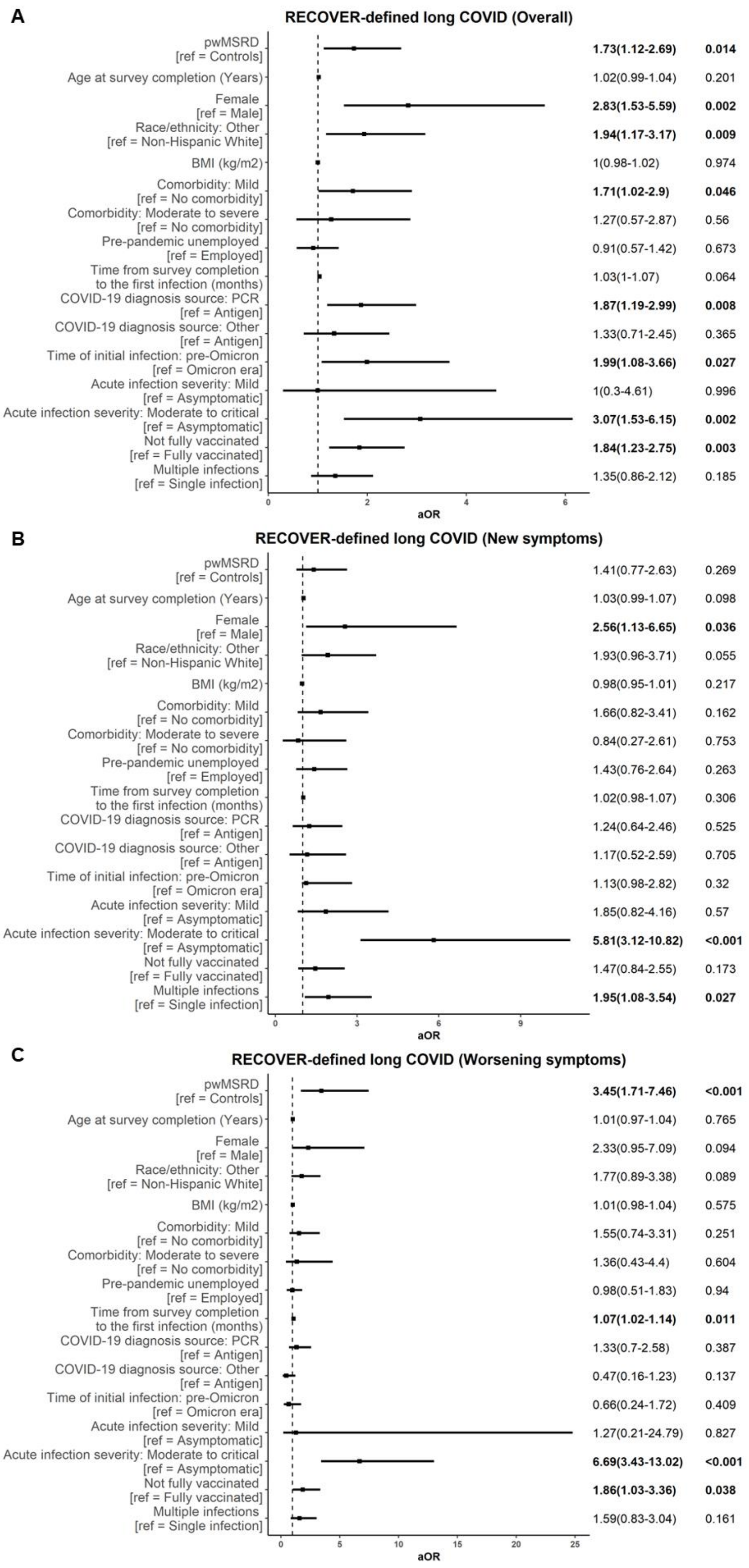
A
RECOVER-defined long COVID (Overall)
pwMSRD [ref = Controls] 1.73(1.12-2.69) 0.014
Age at survey completion (Years) 1.02(0.99-1.04) 0.201
Female [ref = Male] 2.83(1.53-5.59) 0.002
Race/ethnicity: Other [ref = Non-Hispanic White] 1.94(1.17-3.17) 0.009
BMI (kg/m2) 1(0.98-1.02) 0.974
Comorbidity: Mild [ref = No comorbidity] 1.71(1.02-2.9) 0.046
Comorbidity: Moderate to severe [ref = No comorbidity] 1.27(0.57-2.87) 0.56
Pre-pandemic unemployed [ref = Employed] 0.91(0.57-1.42) 0.673
Time from survey completion to the first infection (months) 1.03(1-1.07) 0.064
COVID-19 diagnosis source: PCR [ref = Antigen] 1.87(1.19-2.99) 0.008
COVID-19 diagnosis source: Other [ref = Antigen] 1.33(0.71-2.45) 0.365
Time of initial infection: pre-Omicron [ref = Omicron era] 1.99(1.08-3.66) 0.027
Acute infection severity: Mild [ref = Asymptomatic] 1(0.3-4.61) 0.996
Acute infection severity: Moderate to critical [ref = Asymptomatic] 3.07(1.53-6.15) 0.002
Not fully vaccinated [ref = Fully vaccinated] 1.84(1.23-2.75) 0.003
Multiple infections [ref = Single infection] 1.35(0.86-2.12) 0.185
0 2 4 6
aOR
B
RECOVER-defined long COVID (New symptoms)
pwMSRD [ref = Controls] 1.41(0.77-2.63) 0.269
Age at survey completion (Years) 1.03(0.99-1.07) 0.098
Female [ref = Male] 2.56(1.13-6.65) 0.036
Race/ethnicity: Other [ref = Non-Hispanic White] 1.93(0.96-3.71) 0.055
BMI (kg/m2) 0.98(0.95-1.01) 0.217
Comorbidity: Mild [ref = No comorbidity] 1.66(0.82-3.41) 0.162
Comorbidity: Moderate to severe [ref = No comorbidity] 0.84(0.27-2.61) 0.753
Pre-pandemic unemployed [ref = Employed] 1.43(0.76-2.64) 0.263
Time from survey completion to the first infection (months) 1.02(0.98-1.07) 0.306
COVID-19 diagnosis source: PCR [ref = Antigen] 1.24(0.64-2.46) 0.525
COVID-19 diagnosis source: Other [ref = Antigen] 1.17(0.52-2.59) 0.705
Time of initial infection: pre-Omicron [ref = Omicron era] 1.13(0.98-2.82) 0.32
Acute infection severity: Mild [ref = Asymptomatic] 1.85(0.82-4.16) 0.57
Acute infection severity: Moderate to critical [ref = Asymptomatic] 5.81(3.12-10.82) <0.001
Not fully vaccinated [ref = Fully vaccinated] 1.47(0.84-2.55) 0.173
Multiple infections [ref = Single infection] 1.95(1.08-3.54) 0.027
0 3 6 9
aOR
C
RECOVER-defined long COVID (Worsening symptoms)
pwMSRD [ref = Controls] 3.45(1.71-7.46) <0.001
Age at survey completion (Years) 1.01(0.97-1.04) 0.765
Female [ref = Male] 2.33(0.95-7.09) 0.094
Race/ethnicity: Other [ref = Non-Hispanic White] 1.77(0.89-3.38) 0.089
BMI (kg/m2) 1.01(0.98-1.04) 0.575
Comorbidity: Mild [ref = No comorbidity] 1.55(0.74-3.31) 0.251
Comorbidity: Moderate to severe [ref = No comorbidity] 1.36(0.43-4.4) 0.604
Pre-pandemic unemployed [ref = Employed] 0.98(0.51-1.83) 0.94
Time from survey completion to the first infection (months) 1.07(1.02-1.14) 0.011
COVID-19 diagnosis source: PCR [ref = Antigen] 1.33(0.7-2.58) 0.387
COVID-19 diagnosis source: Other [ref = Antigen] 0.47(0.16-1.23) 0.137
Time of initial infection: pre-Omicron [ref = Omicron era] 0.66(0.24-1.72) 0.409
Acute infection severity: Mild [ref = Asymptomatic] 1.27(0.21-24.79) 0.827
Acute infection severity: Moderate to critical [ref = Asymptomatic] 6.69(3.43-13.02) <0.001
Not fully vaccinated [ref = Fully vaccinated] 1.86(1.03-3.36) 0.038
Multiple infections [ref = Single infection] 1.59(0.83-3.04) 0.161
0 5 10 15 20 25
aOR

**eFigure 9. Multivariable linear regression for PROMIS physical function (left), cognitive function (middle), and depression (right).**

Model 1, RECOVER-defined long COVID overall as the main exposure; Model 2, RECOVER-defined long COVID on new symptoms as the main exposure; Model 3, RECOVER-defined long COVID on worsening symptoms as the main exposure.

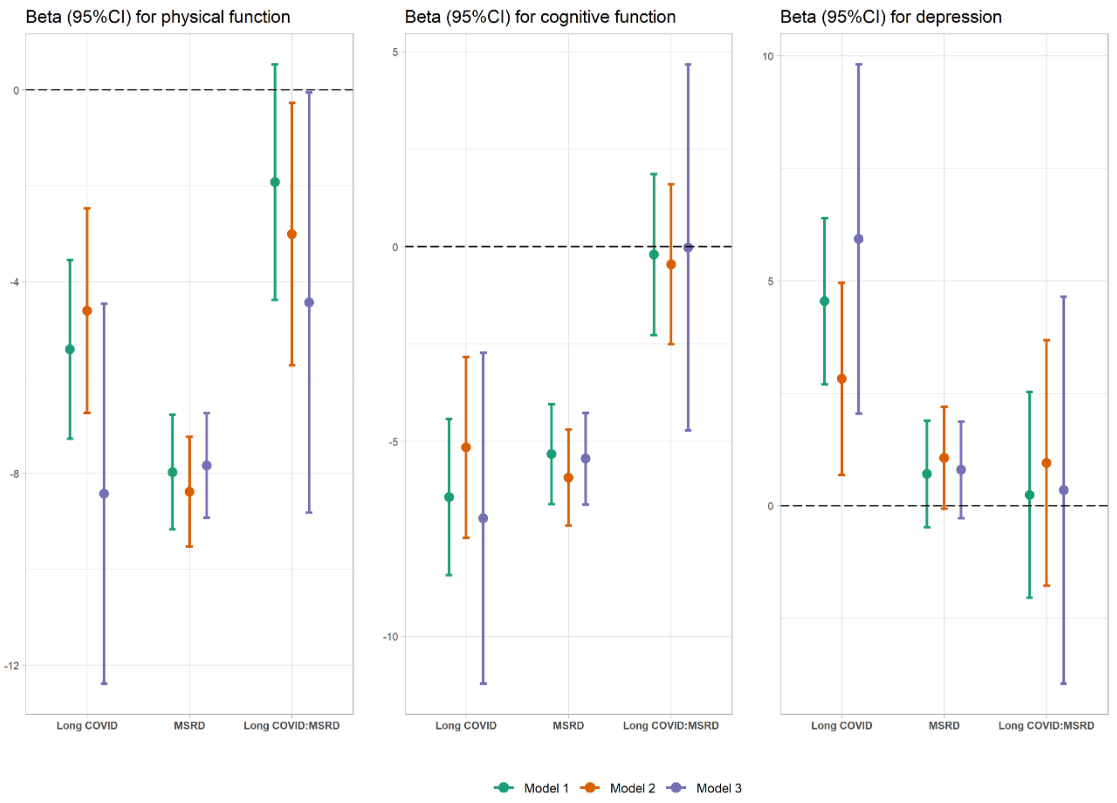
